# Global trends in air travel: implications for connectivity and resilience to infectious disease threats

**DOI:** 10.1101/2020.03.29.20046904

**Authors:** Ashleigh R. Tuite, Deepit Bhatia, Rahim Moineddin, Isaac I. Bogoch, Alexander G. Watts, Kamran Khan

## Abstract

**Background:** Increased connectivity via air travel can facilitate the geographic spread of infectious diseases. The number of travelers alone does not explain risk; passenger origin and destination will also influence risk of disease introduction and spread. We described trends in international air passenger numbers and connectivity between countries with different capacities to detect and respond to infectious disease threats.

**Methods:** We used the Fragile States Index (FSI) as an annual measure of country-level resilience and capacity to respond to infectious disease events. Countries are categorized as: Sustainable, Stable, Warning, or Alert, in order of increasing fragility. We included data for 177 sovereign states for the years 2007 to 2016. Annual inbound and outbound international air passengers for each country were obtained for the same time period. We examined trends in FSI score, trends in worldwide air travel, and the association between a state’s FSI score and air travel.

**Results:** Among countries included in the FSI rankings, the total number of passengers increased from 0.791 billion to 1.28 billion between 2007 and 2016. Increasing fragility was associated with a decrease in travel volumes, with a 2.9% (95% CI: 2.3-3.5%) reduction in passengers per 1-unit increase in FSI score. Overall, travel between countries of different FSI categories either increased or remained stable. The greatest increase was observed for travel to Warning countries from Warning countries, with an annual increase of 8,967,623 passengers (95%CI: 6,546,494 to 11,388,753) over the study period.

**Conclusions:** The world’s connectivity via air travel has increased dramatically over the past decade. There has been notable growth in travel from Warning and Stable countries, which comprise more than three-quarters of international air travel passengers. These countries may have suboptimal capacity to detect and respond to infectious disease threats that emerge within their borders.

## INTRODUCTION

The global population recently surpassed 7.6 billion people and is expected to reach 9.8 billion by 2050.^1^ In tandem, transportation networks have expanded and evolved to satisfy the growing social and economic demands for global connectivity.^2^ Today, commercial air travel is the conduit for approximately 3.5 billion trips annually, of which over 40% are international.^3^ We live in an increasingly connected world, with more people traveling further distances than prior generations and decreased times required to travel these increased distances.^4^ While this interconnectedness is a defining feature of globalization and has produced tremendous benefits for humankind, it has also facilitated the geographic dispersion of infectious diseases.^5^ Although travel has always been associated with the introduction of pathogens to new environments and populations, the speed with which these new introductions occurs has been enhanced as population mobility increases. In the 1800s, the slow march of the second cholera pandemic could be observed to follow trade and military campaign routes out of India to Central Asia, the Middle East, Europe, and eventually North America, over the course of years.^5^ Today, by contrast, infectious diseases can traverse the globe in less than a day.^6-8^ With improvements in the availability of and access to various modes of transportation over the past several decades and the resultant decreases in time required to reach destinations, an outbreak in an isolated location can pose an international threat.^5^

It is generally assumed that increased numbers of international travelers will increase global vulnerability to infectious diseases, by enhancing the potential for geographic spread. However, increased travel volume alone does not capture another important feature of travel trends – connectivity between countries with differential capacities to detect and respond to infectious disease threats. Increased travel between two countries with strong health care and public health systems will likely have very different implications for global health security than increased travel between countries with less developed infrastructure or countries with disparities in their capacities to respond to public health threats. For instance, an increase in travelers to more vulnerable countries may increase the likelihood of exportation of cases to other countries, thereby increasing the rapidity of global transmission. However, if the countries to which the disease is exported have suitable health capacity, the risk of establishment and ongoing transmission may be less of a concern,^9^ albeit non-negligible.^10^ Increased connectivity also increases the likelihood of introduction of pathogens via infected tourists to less resilient countries, where disease establishment is a risk.^11^

To better understand the impact of globalization on our ability to mitigate infectious disease threats, we sought to describe trends in air passenger numbers over a 10-year period (2007-2016) and investigate if connectivity between countries with different levels of resilience has changed over this time period.

## METHODS

### Data sources

There are various indices that have been developed to measure the resilience and capacity of countries to respond to various social, economic, and political pressures, including public health emergencies.^12, 13^ The Fragile States Index is one such measure that has been calculated annually since 2005, capturing changes in countries’ resilience and ability to respond to economic and political pressures over time.^14^ We used this index as a measure of capacity to respond to infectious disease threats. As of 2016, the Index included 178 sovereign states. Countries are ranked on 12 indicators. Each indicator is scored on a scale of 0 to 10, with 0 being the most stable and 10 the most fragile, producing an overall score ranging from 0 to 120. Of the 12 indicators, 4 are categorized as social, 2 as economic, and 6 as political. Based on overall scores, states are assigned to one of four categories (in order of increasing fragility): Sustainable (0-29.9), Stable (30-59.9), Warning (60-89.9), and Alert (90-120). States in the Stable, Warning, and Alert categories are considered vulnerable to failure. We obtained FSI numeric scores and categorical rankings for all available countries for the years 2007 to 2016.

To quantify international population mobility, we analyzed worldwide airline ticket sales data from the International Air Transport Association (IATA) for the time period covering January 1, 2007 to December 31, 2016. IATA captures passenger-level data on an estimated 90% of all scheduled commercial air travel bookings worldwide (domestic and international) and imputes values for the remaining 10% of passenger trips using a proprietary algorithm.^3^ We restricted the analysis to international passengers only. For each country, we obtained annual inbound and outbound passenger numbers.

### Analysis overview

We examined trends in FSI score, trends in worldwide air travel, and the association between a state’s FSI score and air travel. Travel between countries included in the FSI rankings represented 95% (1.28 billion out of 1.35 billion) of all international trips in 2016. We excluded South Sudan from these analyses, as FSI data were only available from 2012 onwards. South Sudan is classified as an Alert country and travel to and from this country accounted for less than 0.03% of annual passenger flows. Analyses were conducted using either inbound or outbound passenger numbers, but are presented for outbound passenger numbers only, as the inbound and outbound passenger volumes did not differ significantly (paired T-test, p=0.62).

### Assessing trends in national resilience

We assessed changes in FSI over time using a random intercept and random slope mixed model to account for changes within each country by year, with FSI value as the dependent variable and time as the independent variable.

### Assessing trends in connectivity based on national resilience

We evaluated trends in outbound passenger numbers over time for each FSI category, using linear regression models with travel volume as the dependent variable and time as the independent variable. To quantify the association between FSI and travel volume, we used a model with repeated yearly measures at the country level, using an autoregressive correlation matrix. The dependent variable was travel volume, while the independent variables were time and FSI value. To quantify the change in travel volumes over time at the individual country level, we used a mixed model with random intercept and slope terms that differed at the country level, with an unstructured covariance matrix.

To examine how travel between countries belonging to different FSI categories has changed over time, we calculated annual absolute number of outbound passengers between origin-destination FSI category combinations (e.g., total passengers from Alert to Sustainable countries). We also determined the proportionate amount of outbound travel to each destination FSI category for a given origin FSI category. To quantify the change in travel between FSI category combinations, we used a mixed model with random slope and intercept for each origin-destination category pair (such as “Alert to Sustainable”). Using this model, we obtained the yearly change in passenger volume between each category. We log transformed the number of travelers to stabilize the variance and eliminate the skewness. Analyses were conducted in SAS Enterprise Guide 7.1 (SAS Institute, Cary NC) and R.^15^

## RESULTS

### Trends in international population mobility

Among countries included in the FSI rankings, the total number of passengers increased from 0.791 billion to 1.28 billion, representing a greater than 60% increase between 2007 and 2016. Total distance traveled increased from 5.35 trillion to 8.64 trillion kilometers over this time period.

### Trends in national resilience

In 2007 and 2016, 72% and 69% of countries, respectively, were categorized as Warning or Alert, indicating that ‘significant parts of their societies and institutions were vulnerable to failure’ ^14^ (**Figure 1**). Throughout this time period, the largest proportion of countries were assigned to the Warning category (**Table 1**), representing the second most vulnerable ranking.

**Table 1.** Annual distribution of Fragile State Index categories for 177 countries, 2007-2016.

| Year | Percent (n) |  |  |  |
| --- | --- | --- | --- | --- |
|  | Sustainable | Stable | Warning | Alert |
| 2007 | 8.5 (15) | 19.8 (35) | 52.0 (92) | 19.8 (35) |
| 2008 | 7.3 (13) | 18.6 (33) | 53.1 (94) | 20.9 (37) |
| 2009 | 7.3 (13) | 19.8 (35) | 52.0 (92) | 20.9 (37) |
| 2010 | 6.8 (12) | 23.2 (41) | 50.3 (89) | 19.8 (35) |
| 2011 | 7.3 (13) | 22.0 (39) | 52.0 (92) | 18.6 (33) |
| 2012 | 7.9 (14) | 21.5 (38) | 51.4 (91) | 19.2 (34) |
| 2013 | 7.3 (13) | 22.0 (39) | 52.0 (92) | 18.6 (33) |
| 2014 | 8.5 (15) | 21.5 (38) | 49.7 (88) | 20.3 (36) |
| 2015 | 9.0 (16) | 20.9 (37) | 49.2 (87) | 20.9 (37) |
| 2016 | 8.5 (15) | 22.0 (39) | 50.3 (89) | 19.2 (34) |

**Figure 1.**
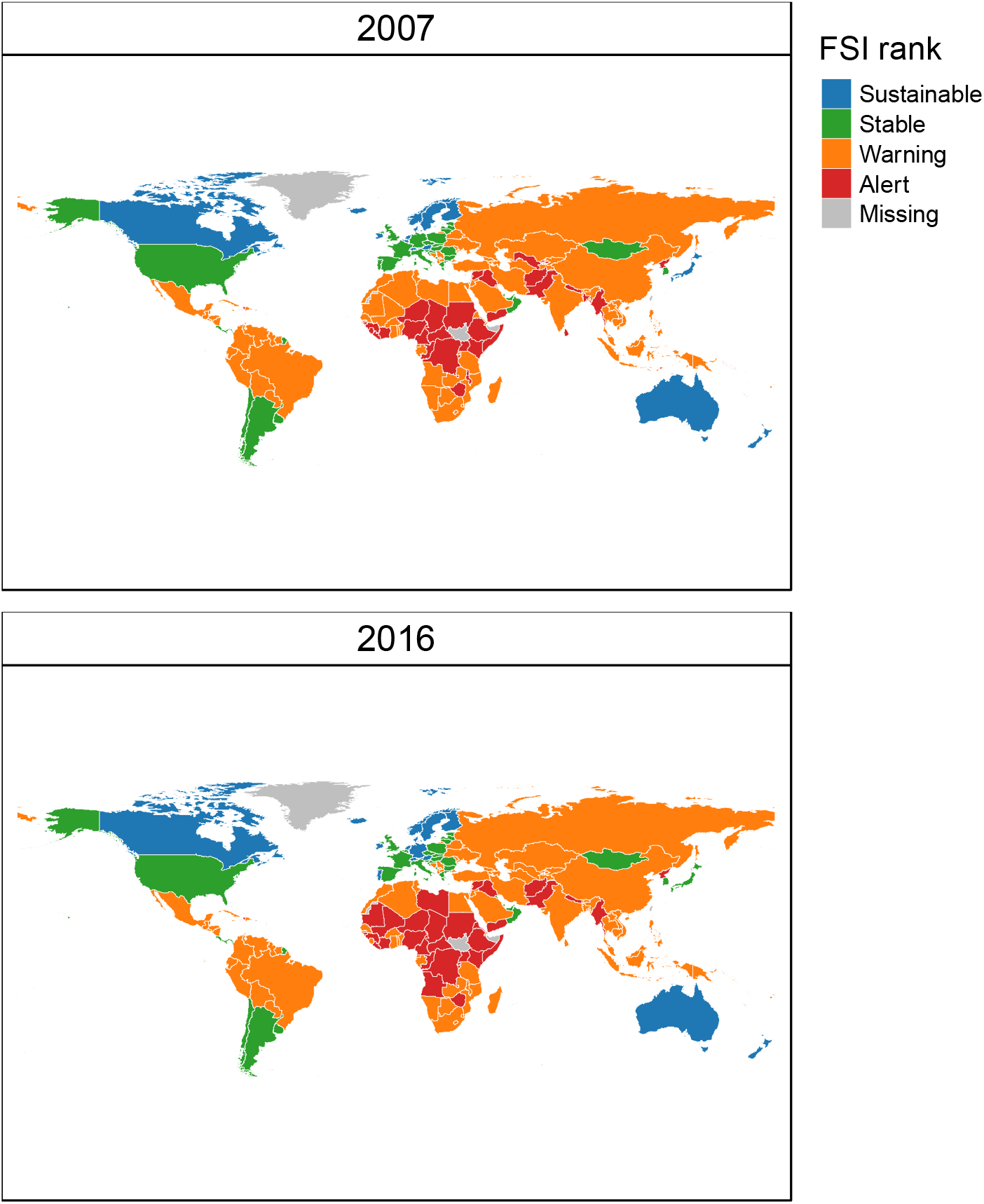
Country Fragile State Index ranks, 2007 and 2016. FSI ranks are shown for the 177 countries with data for both years. Note that South Sudan is excluded as there was no rank prior to 2012, but had an FSI rank of ‘Alert’ in 2016. FSI ranks correspond to the following FSI scores, with a higher score indicating a more fragile state: Sustainable, 0-29.9; Stable, 30-59.9; Warning, 60-89.9; Alert, 90-120.

There was a trend of decreased fragility over the 10-year time period, with FSI values decreasing by 0.23 (95% CI: −0.34, −0.11) points per year. Comparing 2007 and 2016, a total of 23 countries (13%) had a change in FSI category, with the remaining 154 countries remaining in the same category at both time points. Although the number of countries with a change in category was relatively small, many countries did experience changes in their numeric FSI scores. Between 2007 and 2016, 44 countries (25%) had an increase in their FSI score of 5% or more, indicating increasing fragility, with Libya (+26.3 points), Syria (+20.5 points), and Mali (+17.3 points) experiencing the largest increases in fragility. Over this same period, 70 countries (40%) had a decrease in their FSI score of 5% or more, indicating decreasing fragility. The largest changes were for Cuba (−14.0 points), Moldova (−13.7 points), and Belarus (−12.0 points).

### Trends in connectivity based on national resilience

Between 2007 and 2016 there was an absolute increase in outbound passengers for all FSI categories (**Figure 2**). The largest number of passengers travelled from Stable countries, which experienced a non-significant increase of 298,554 passengers per year (95%CI: −481,794 to 1,078,903). The Alert category had a total increase of 47,022 passengers per year (95%CI: 523 to 93,520), the Warning category had a total increase of 260,420 passengers per year (95%CI: 123,645 to 397,195), and the Sustainable category had a total increase of 872,469 passengers per year (95%CI: 286,761 to 1,458,178). We also observed a statistically significant relationship between FSI value and travel volumes, independent of the effect of time (**Figure 2**). Each 1-unit increase in FSI score (increasing fragility) in a country was associated with a decrease of 62,595 passengers (95% CI: 38,358 to 86,832, **Table S1**). This is equivalent to a 2.9% (95% CI: 2.3-3.5%) reduction in passengers per 1-unit increase in FSI score (**Table S2**).

**Figure 2.**
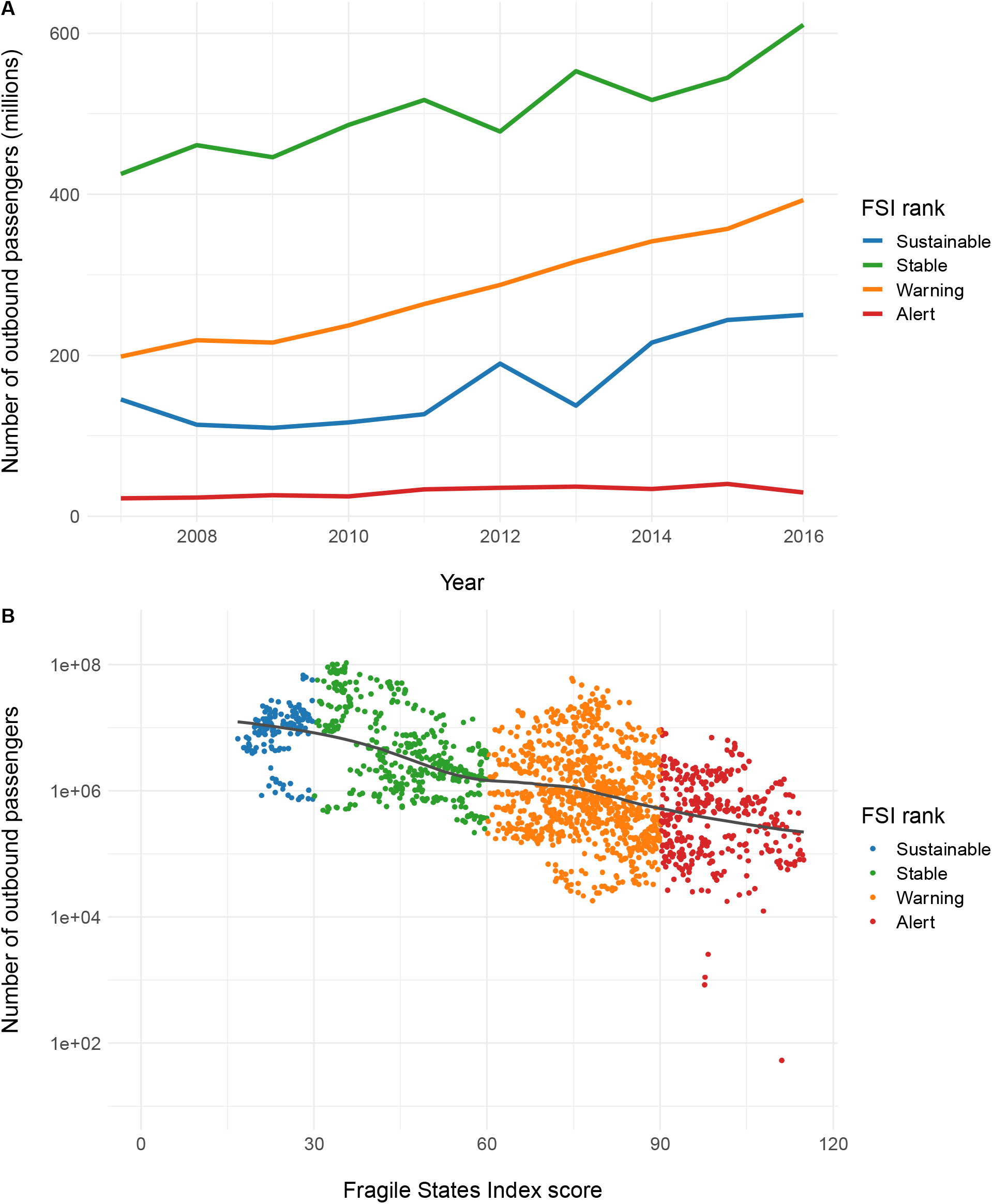
Relationship between Fragile State Index score and outbound passenger numbers. (A) Changes in outbound passenger numbers over time, by Fragile State Index rank. (B) Relation between FSI score and number of outbound passengers. Note that outbound passengers are plotted on a logarithmic scale for (B). Results are shown for all countries for the time period 2007 to 2016.

The annual percent change in outbound travel varied considerably across countries (**Figure 3**). Only Syria and Yemen experienced statistically significant decreases in outbound passengers over time. Fifty-five percent of countries had a statistically significant increase in the annual percentage of outbound passengers over time, with the largest percent changes occurring in Alert countries (North Korea, Iraq, and Myanmar).

**Figure 3.**
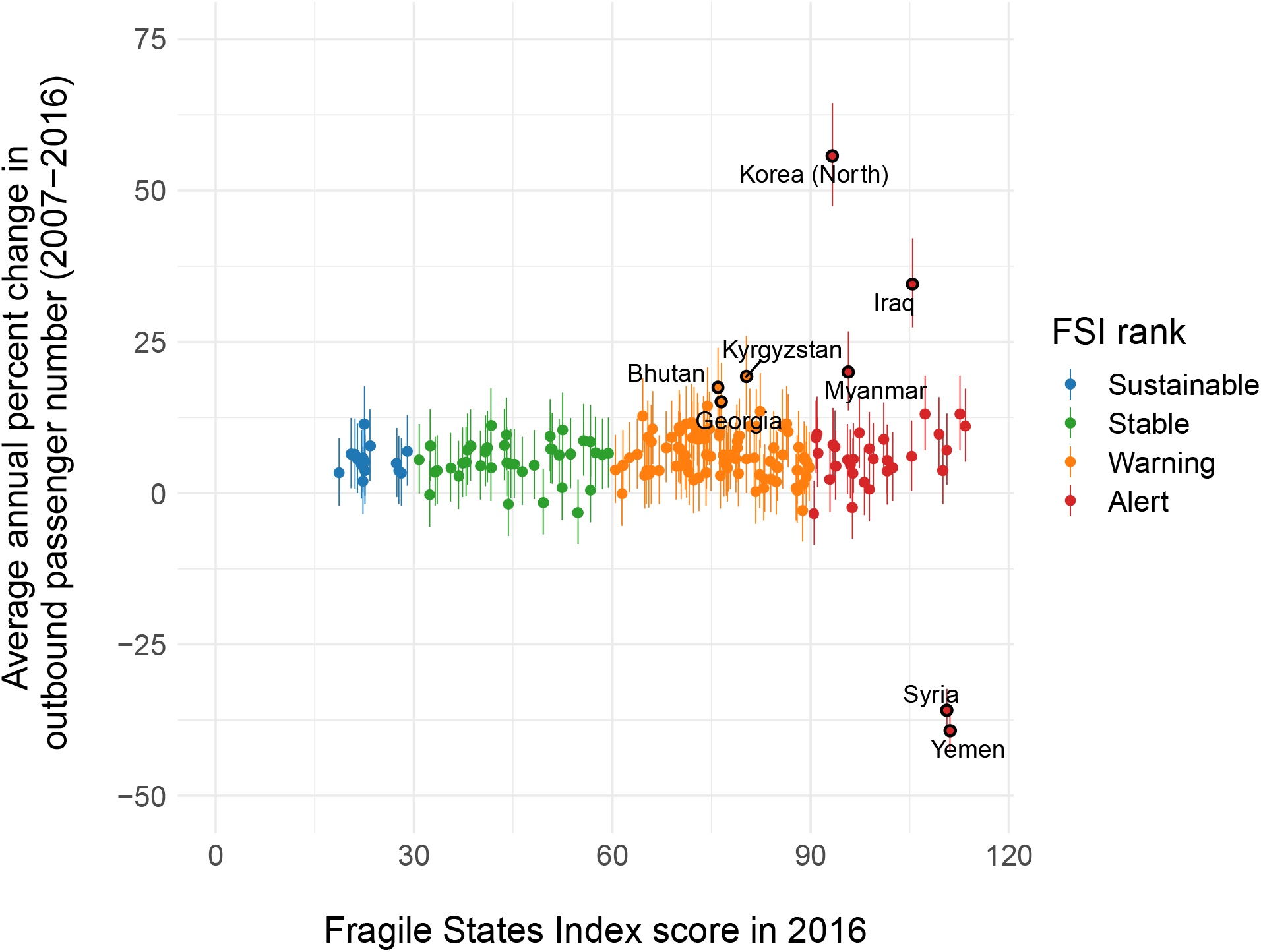
Annual percent change in outbound passenger numbers for each country (n=177) included in the FSI rankings for the period 2007 to 2016. The x-axis shows country FSI scores in 2016 and colors indicate the ranking associated with the score. Countries with annual changes in passenger volumes of +/-15% are labeled and outlined in black. Alert represents the most fragile states and Sustainable the least fragile.

An example of how total passenger numbers and connectivity between countries belonging to different FSI categories has changed between 2007 and 2016 is presented in **Figure 4**. The proportion of outbound passengers departing from Warning and Alert countries to other fragile states increased over time (**Figure 5**). Passengers from Sustainable and Stable countries traveled most frequently to countries within the same categories. Overall, travel between countries of different FSI categories either increased or remained stable. The origin-destination pair with the greatest increase was travel to Warning countries from Warning countries, with an annual increase of 8,967,623 passengers (95%CI: 6,546,494 to 11,388,753) (**Figure 5**). There were no statistically significant increases in travel to or from Alert category countries.

**Figure 4.**
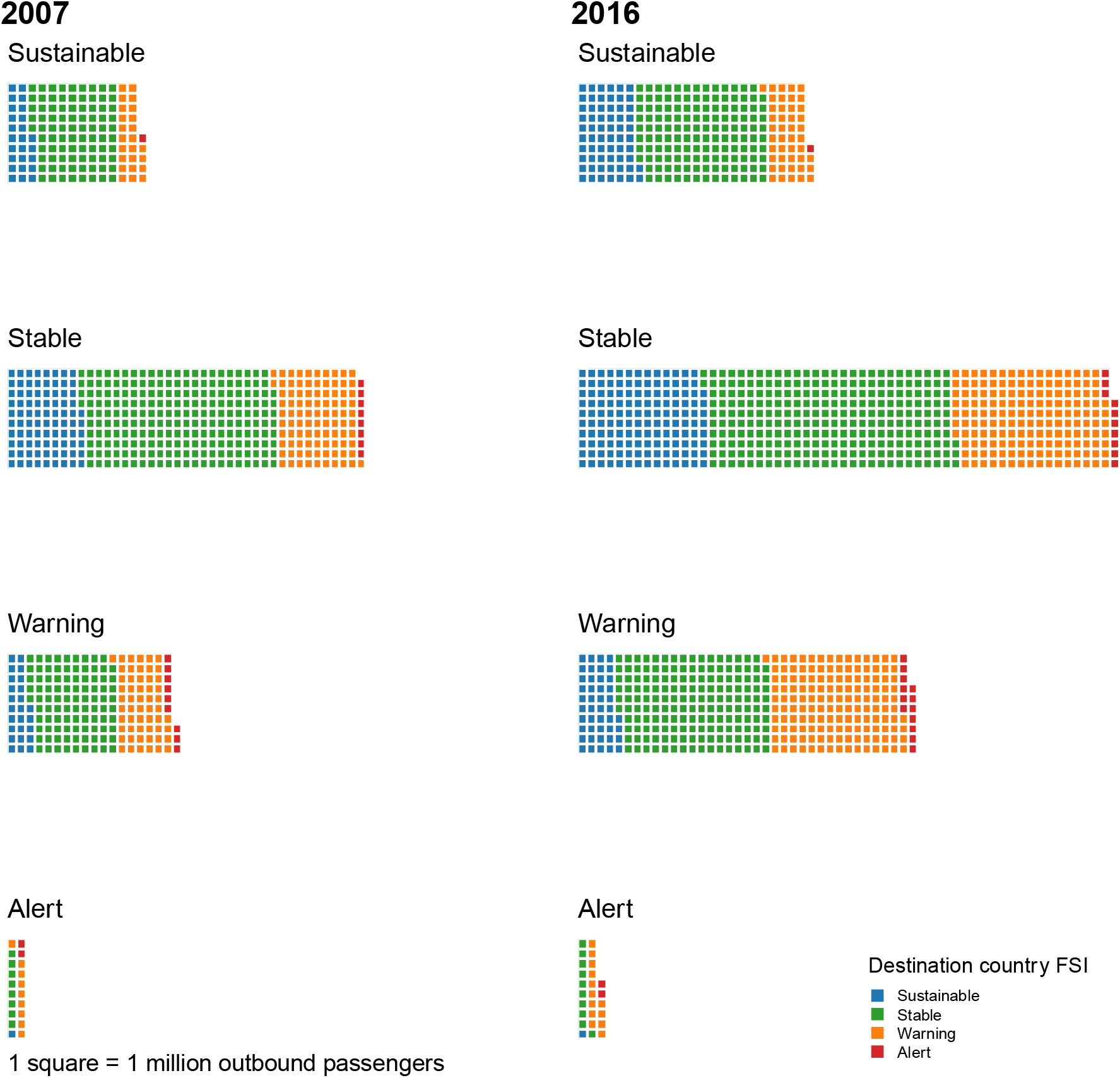
Comparison of passenger trends for the years 2007 and 2016. For each year, results are presented by origin country FSI rank. Each square represents 1 million outbound passengers. The number of squares represents the number of outbound passengers from countries of a given FSI rank. Squares are coloured by the FSI rank of the destination country. Alert represents the most fragile states and Sustainable the least fragile.

**Figure 5.**
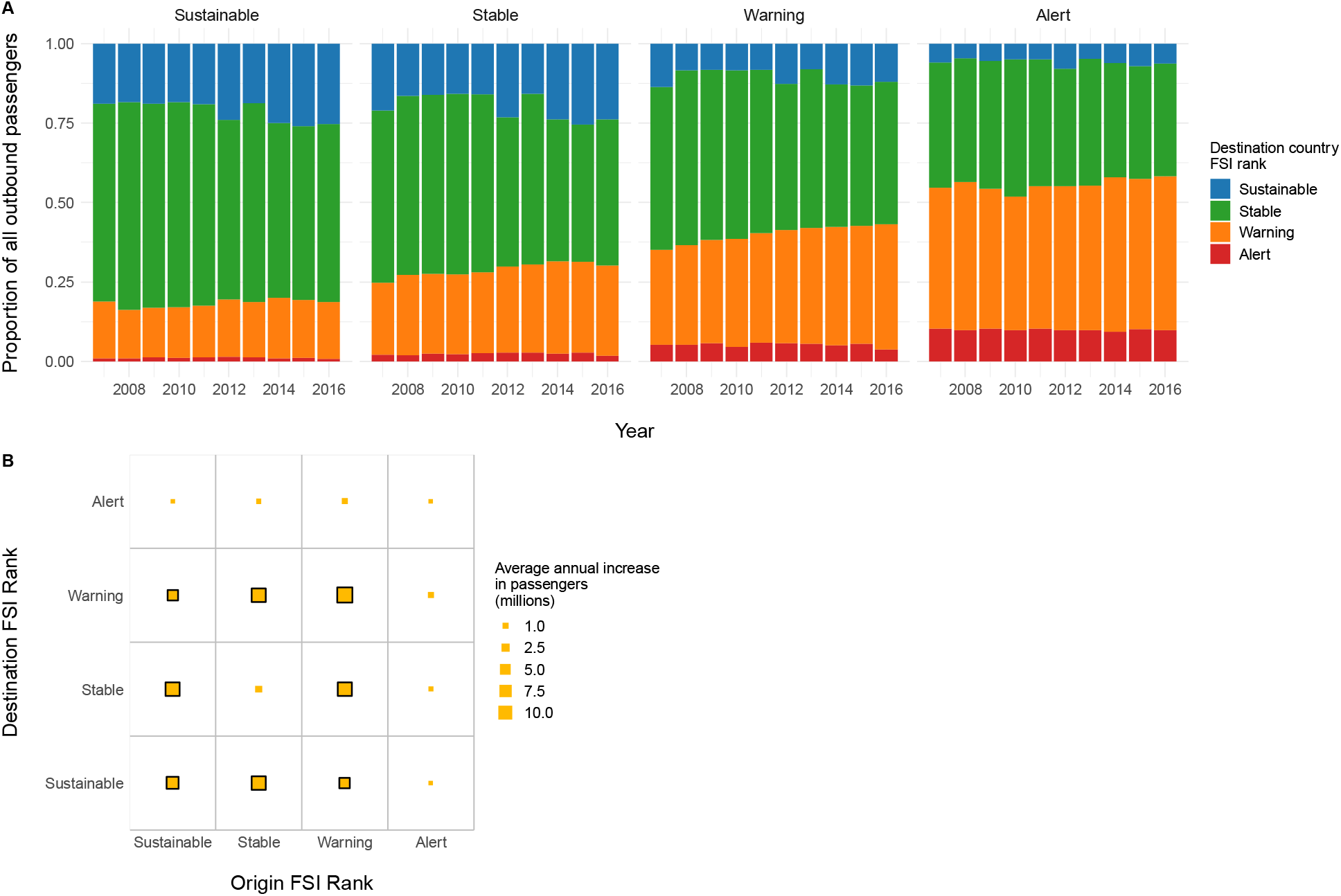
Changes in connectivity between countries with different FSI rankings, 2007-2016. (A) The proportion of outbound passengers from countries of a given FSI rank arriving to countries with different FSI ranks is shown for each year. Results are presented by FSI rank of the origin countries for each year. (B) Average annual growth in outbound passengers from origin to destination countries of different FSI ranks. Box sizes represent average annual increases in passenger volumes (in millions) over the 10-year time period between each pair of FSI categories. Origin-destination pairs with statistically significant increases (p<0.05) are indicated by black outlines around boxes.

## DISCUSSION

The world’s connectivity via air travel has increased dramatically over the past decade, potentially increasing the vulnerability of billions of people across the world to existing and novel pathogens without a corresponding increase in coping capacity.^5, 16, 17^ Using the Fragile States Index and airline passenger data, we show that although the number of passengers traveling internationally is increasing, this growth is not uniform across all FSI categories. In particular, although there has been an increase in travel from the most vulnerable Alert countries, the overall amount of travel from these countries is small, relative to overall passenger volumes. There has been notable growth in travel from Warning and Stable countries, which comprise more than three-quarters of international air travel passengers, and which may have suboptimal capacity to detect and respond to infectious disease threats that emerge within their borders.

The importance of less resilient countries as sources of public health threats is highlighted by an analysis of the 46 countries with FSI scores that had one or more disease outbreaks reported by the World Health Organization in 2016 ^18^: 22% were Alert countries and 48% were Warning countries. Only 7% were ranked as Sustainable.

The large increases in reciprocal travel between countries belonging to the Warning category is of concern due to the potential for greater exposure to and possible spread of pathogens across regions with less developed health capacity.^5, 19-21^ Furthermore, as the FSI is a composite index aggregating indicators across different sectors (social, economic, and political), countries with the same FSI value could have vulnerabilities across these different dimensions. Mixing of populations coming from comparably Fragile states, but with different vulnerabilities, could potentially result in synergies that increase the magnitude and impact of infectious disease events.

Despite the robust overall trend of increasing air travel over time, we also observe a relation between increasing FSI (i.e., increasing vulnerability) and reductions in air travel, suggesting that changes in individual country’s vulnerability does have repercussions for air travel connectivity.

In addition to the population health implications of changes in the destination and volume of international travellers, our findings are also important for individual traveler health. With an increase in the number of visitors to Alert and Warning countries, the number of travel-related exported infectious cases with epidemic potential may increase. This, in turn, may affect tourist industries, resulting in human and economic losses.^22^

Quantifying a country’s ability to detect and respond to infectious disease threats is challenging, as there is a constellation of complex and evolving socioeconomic, political, and environmental factors that contribute to that ability. The FSI and other indices attempt to distill these multidimensional factors into a single metric, which of necessity will be a relatively crude approximation. Unlike other such indices,^12, 13^ the FSI is unique because it captures year-to-year changes, allowing us compare changes in each country’s Fragility over time.^14^ The use of the FSI does come with limitations. The data are also not available for all countries, limiting the scope of the analyses. The classification of countries into 4 categories based on numeric scores, while useful for communicating and understanding the risk levels, may reduce power to detect trends in the data. To address this, we used the continuous FSI score when possible. The data are only available at the country level, which does not allow for analyses at a finer geographic scale. A key assumption in this analysis is that passenger volumes are a measure of importation pressure, with humans acting as the ‘vectors’ of disease.^23^ This does not account for important differences in population vulnerability, such as vaccination status, pre-existing health conditions, specific locations where tourists may be likely to visit, and reasons for traveling.

In summary, there is an association between changes in the coping capacity of a country to manage outbreaks, and changes in outgoing passenger volumes. Air travel will likely continue to increase, outpacing the improvements in our ability to prevent, detect and control epidemics, especially in resource limited settings. While air travel remains a safe and rapid means of connecting people across the world, the impact of even one exported case can be catastrophic, emphasizing the importance of strengthening global health capacity and security.

## Data Availability

Fragile State Index data are publicly available. The air travel data are proprietary.

## Supplementary Tables

**Table S1.** Relation between Fragile States Index score and outbound travel volumes, accounting for time. Coefficients for year and FSI score represent changes in travel volumes associated with a one-year increase or one-unit change in FSI score, respectively. Note that an increase in FSI score represents an increase in Fragility.

| Effect | Estimate | Standard Error | DF | t Value | Pr > \|t\| |
| --- | --- | --- | --- | --- | --- |
| Intercept | 8898855 | 1300040 | 176 | 6.85 | <.0001 |
| Year | 300254 | 22310 | 1591 | 13.46 | <.0001 |
| FSI score | -62595 | 12366 | 1591 | -5.06 | <.0001 |

**Table S2.** Relation between Fragile States Index score and the percent change in outbound travel volumes, accounting for time. Coefficients for year and FSI score represent changes in travel volumes associated with a one-year increase or one-unit change in FSI score, respectively.

| Effect | Percent change | 95% lower confidence limit | 95% upper confidence limit |
| --- | --- | --- | --- |
| Year | 5.093 | 3.676 | 6.530 |
| FSI score | -2.895 | -3.501 | -2.286 |

